# Estimating dengue force of infection from age-stratified surveillance data in Java, Indonesia

**DOI:** 10.1101/2025.05.16.25327741

**Authors:** Bimandra A Djaafara, Iqbal RF Elyazar, Asik Surya, Fadjar SM Silalahi, Agus Handito, Desfalina Aryani, Mushtofa Kamal, Dyana Gunawan, Hipokrates, Anzala Khoirun Nisa, Edi Prianto, Iriani Samad, Agus Sugiarto, Burhannudin Thohir, Hannah E Clapham, Swapnil Mishra

## Abstract

Targeted dengue interventions require reliable estimates of transmission intensity and population immunity at the local level. The force of infection (FOI) provides an objective measure of transmission intensity, but its estimation traditionally relies on resource-intensive seroprevalence surveys. We developed a hierarchical extension of existing catalytic models to estimate FOI using routine age-stratified surveillance data, allowing partial pooling of information across districts within provinces. We applied this approach to dengue surveillance data from Jakarta and West Java provinces, Indonesia, and compared it with non-hierarchical implementations. Both hierarchical and non-hierarchical approaches produced FOI estimates consistent with 2014 seroprevalence data. The hierarchical framework provided more robust estimates through partial pooling under varied data availability scenarios but showed sensitivity to age-stratification choices and could miss district-specific patterns when local epidemiology differed from regional trends. Model comparison using Expected Log Pointwise Predictive Density showed that accounting for overdispersion through negative binomial likelihood improved model performance regardless of hierarchical structure. Our analysis showed distinct patterns in reporting parameters between provinces, with Jakarta showing higher reporting rates despite lower FOI estimates than West Java. Implementation of the hierarchical framework requires understanding of local dengue epidemiology, as clustering districts with different epidemiological profiles could produce inaccurate estimates.

## Introduction

Dengue virus transmission continues to pose a significant public health challenge across tropical and subtropical regions, with Indonesia experiencing one of the highest burdens in Southeast Asia [1]. In Indonesia, dengue is also the most common cause of acute febrile illness requiring hospitalisation [2]. In 2021, the Ministry of Health (MoH) of the Republic of Indonesia launched its National Strategic Plan (NSP) for the dengue control programme to achieve zero dengue deaths by 2030 and ensure that at least 90% of cities and districts maintain a dengue incidence rate below 49 per 100,000 and case fatality ratio below 0.5% by 2025 [3,4]. With the recent availability and rollout of novel interventions for dengue, such as Wolbachia-treated mosquitoes and vaccines [5], Indonesia has potentially effective tools to control dengue. However, nationwide implementation of these interventions requires substantial resources and may not be immediately feasible across all regions. Moreover, dengue transmission intensity varies substantially across Indonesia’s geography due to differences in climate, urbanisation, and socioeconomic factors [6,7], suggesting that a uniform nationwide approach may not be optimal. Therefore, strategic targeting of interventions is needed for optimal resource allocations to achieve maximum impact.

A fundamental challenge in this strategic planning lies in transforming routinely collected surveillance data (mainly through passive surveillance) into actionable evidence for control interventions. While disease surveillance systems typically rely on analysing aggregate case counts to stratify population risks of dengue, raw case counts or incidence rates can be misleading indicators of true disease burden, especially for dengue, where reporting variations may yield from surveillance system design (case definition, laboratory confirmation, inpatient or outpatient, etc.) and surveillance implementation (treatment-seeking behaviour, paper or electronic forms, etc.) [8,9]. In Indonesia specifically, the surveillance system captures only dengue haemorrhagic fever (DHF) cases requiring hospitalisation, leading to substantial underreporting of the total dengue burden [10,11]. Additionally, Indonesia has implemented an Early Warning and Response System (EWARS) to capture suspected dengue cases at the primary health care level [12]. The system adopts a syndromic approach to detect potential outbreaks and collects aggregated weekly data from over 10,000 primary health facilities across the country. Surveillance quality also often varies significantly across and within countries, with areas with more complete reporting and better surveillance systems potentially appearing to have higher disease burden simply due to better case detection or reporting [13,14]. Furthermore, temporal changes in surveillance systems, such as improvements in surveillance and reporting over time, can complicate historical comparisons and trend analyses.

The force of infection (FOI), the per capita rate at which susceptible individuals acquire infection, is a metric used for quantifying transmission intensity. Cross-sectional seroprevalence surveys and age-stratified case notification data from routine surveillance systems have been widely used to estimate dengue FOI [15–21]. While seroprevalence surveys provide direct evidence of past infections, they are resource-intensive and often only available at a limited number of locations and time points [15]. Recent studies have demonstrated that catalytic models fitted to age-stratified surveillance data can generate FOI estimates comparable to those derived from seroprevalence surveys [15,16], validating this more readily available data source for transmission intensity estimation. Furthermore, FOI estimates enable public health officials to overcome some limitations of raw surveillance data by providing a standardised measure of transmission that accounts for population immunity, age-specific infection patterns, and reporting variation across locations.

Given Indonesia’s heterogeneous dengue landscape, FOI estimates can provide critical insights for targeted intervention strategies. FOI estimates can also be used further to calculate other key epidemiological metrics, such as the proportion of susceptible populations and the potential burden of unreported infections.

Given the limitations in surveillance data quality and completeness across different regions, improved methodological approaches are needed to generate robust estimates of transmission intensity. We employed hierarchical Bayesian modelling to extend existing catalytic models for dengue age-stratified incidence data, enabling partial pooling of information across districts within provinces. This approach aims to improve FOI estimates, particularly in districts with sparse or noisy data, leveraging information from neighbouring districts within the same region.

Our study focused on provinces and districts in Java Island as a test case for this methodological development. We analysed data from Jakarta province (6 districts) and West Java province (27 districts). We developed a suite of experiments to systematically evaluate where the hierarchical approach provides the greatest benefits and where its advantages may be limited. These experiments included: 1) validation through comparison with existing seroprevalence data in selected districts; 2) examination of sensitivity to different age-stratification schemes and temporal data availability; and 3) assessment of estimates performance against traditional non-hierarchical methods, which includes evaluation of relationships between model parameters to understand potential limitations in parameter identifiability. Through these experiments, we identified specific data contexts and regional characteristics where the Bayesian hierarchical modelling approach may offer substantial improvements in FOI estimation.

## Methods

### Data

We used age-stratified reported dengue incidence data at the district level from two provinces: Jakarta (DKI Jakarta) and West Java (Jawa Barat). For Jakarta data, we obtained the data from the Jakarta Provincial Health Office’s Immunisation and Surveillance section website [22], and the data for West Java were obtained from the arbovirus working group of the Indonesia Ministry of Health. For Jakarta, we analysed yearly reported dengue haemorrhagic fever (DHF) incidence from all hospitals at the district level from 2017-2023 (7 years), stratified into nine age groups: 0-4, 5-9, 10-14, 15-19, 20-44, 45-54, 55-64, 65-74, and ≥75 years old. For West Java, we used yearly reported age-stratified DHF incidence data from all reporting health facilities between 2016-2023 (8 years), stratified into four age groups: 0-4, 5-14, 15-44, and ≥45 years old.

The age-stratified population data were interpolated and extrapolated from the 2010 [23] and 2020 [24] census data. We used linear interpolation and extrapolation of the age-stratified population counts at the district level to estimate the population counts for years between 2010 and 2020, and after 2020, respectively. For 2020, we used population counts from the census results.

### Modelling framework and comparison

We developed and compared four modelling approaches:

1. Hierarchical model with Poisson likelihood
2. Hierarchical model with negative binomial likelihood
3. Non-hierarchical model with Poisson likelihood
4. Non-hierarchical model with negative binomial likelihood

The hierarchical models implemented a partial pooling of information across districts within each province, while the non-hierarchical models fitted data independently for each district. Two different likelihoods were used to assess whether accounting for overdispersion improved model performance.

### Estimating time-varying dengue force of infection (FOI) from age-stratified incidence data

Catalytic models fitted to reported age-stratified incidence data have been developed and used to estimate the dengue FOI (λ) [15,17–19]. We built upon the previous implementations of the catalytic model [15,17–19] and developed a two-level Bayesian hierarchical catalytic model, allowing for information pooling across districts modelled within a province to estimate the time-varying FOI (λ_*t*_), which represents the per capita rate at which susceptible individuals acquire infection in year *t*, at the district level.

Due to the absence of serotype-specific incidence data, we assumed all four circulating dengue serotypes have a similar FOI. The total force infection of all circulating dengue serotypes is written as *4*λ. We assumed the secondary infection is more likely to cause severe dengue, contributing more to the reported incidence than the primary incidence. We also assumed that the contributions of tertiary and quaternary infection to the reported dengue incidence were negligible.

Our model incorporated two key reporting parameters: 1) the overall reporting parameter (ρ), which captures the proportion of all dengue infections detected by the surveillance system, reflecting factors such as healthcare access, diagnostic and surveillance capabilities, and reporting practices; and 2) the relative reporting ratio parameter (γ), which represents the probability of reporting primary infections relative to secondary infections. When γ < *1*, secondary infections are more likely to be reported than primary infections, a pattern typically observed in dengue-endemic settings where secondary infections often produce more severe clinical presentations [25].

We estimated the historical FOI at district *d* (λ_*d*,*H*_), which is defined as yearly FOI for every year before the reported district-level data became available for estimations, and the current FOI at district *d* (λ_*d*,*t*_), which is defined as yearly force infection during the period of years where district-level data became available for estimations in each province. We estimated λ_*d*,*H*_ by implementing a two-phase approach. For the most recent 10 years before the availability of district-level data, we estimated yearly varying FOI (λ_*recent*,*d*,*H*_). For the remaining historical period (up to 80 years in the past), we estimated a single constant FOI (λ_*constant*,*d*,*H*_). In the hierarchical framework, both the recent and constant historical FOI were subject to partial pooling across districts within provinces.

The proportion of the population aged *a* years old susceptible to all four dengue serotypes at year *t* and district *d* is modelled as:

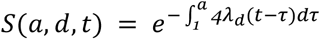

For those aged 0 years old, we assumed full susceptibility to all four dengue serotypes. The proportion of people aged *a* years old in district *d* at year *t* who already experience primary dengue infection (monotypic) is modelled as:

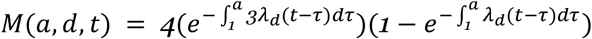

Hence, the incidence rate of primary dengue infections for people aged *a* years old in district *d* at year *t* is modelled as:

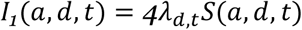

The incidence rate of secondary dengue infections for the same population is:

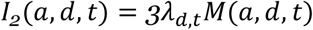

The modelled primary and secondary incidence rates were summarised into the modelled age-stratified reported dengue incidence for model fitting. If we stratified the reported incidence into *G* age groups (*A* = *1*, *2*, *3*, …, *g*), with the lower age bound of the age group *g* as *l*_*g*_ and the upper age bound as *u*_*g*_, the modelled age-stratified reported dengue incidence for age group *g* in district *d* at year *t* is:

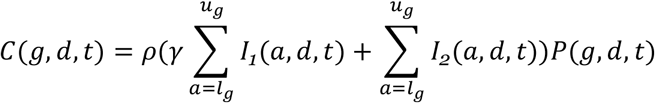

where ρ is the scaling parameter for reporting dengue infection in healthcare facilities, γ is the relative reporting of primary infection compared to secondary infection, and *P*(*g*, *d*, *t*) is the population of *g* age group in district *d* at year *t*.

The modelled age-stratified reported dengue incidence, *C*(*g*, *d*, *t*), were fitted to the reported age-stratified dengue incidence, *R*(*g*, *d*, *t*), assuming Poisson and negative binomial likelihoods in a Bayesian framework:

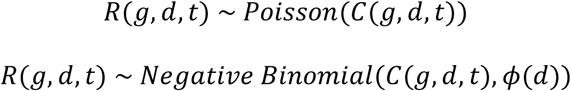

with ϕ(*d*) as the overdispersion parameter at the district level.

For the hierarchical framework, the model fitting processes were done independently in each province for all districts within the province which allows for partial pooling of the parameters. For the non-hierarchical framework, the model fitting processes were done independently in each district. The province- and district-level priors for the parameters of the hierarchical model, and the district-level priors for the parameters of the non-hierarchical model are listed in **the Supplementary Information** (**SI**).

Model validations were done for selected districts where seroprevalence surveys were conducted in 2014 [26]. Modelled estimates of age-specific seroprevalence and overall FOI in those districts were compared to the results from the 2014 study.

Using the estimated parameters from different models, we also simulated some policy-relevant metrics, such as seroprevalence at 9-year-olds, which is used by WHO to define areas with high transmission of dengue [27]. These simulations allowed us to assess how different models might produce different outcomes that may influence relevant policy decisions.

### Estimating dengue FOI from seroprevalence data

Estimates of FOI from seroprevalence data were used for model validations in several districts where surveys were conducted in 2014 [26]. We used a commonly used catalytic model framework to estimate dengue FOI from seroprevalence data. The framework assumes time-constant FOI (4λ) with no waning immunity [20,21].

The proportion of people in age group *g* who were seropositive (*Z*(*g*)) is given by the equation:

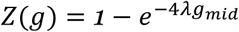

where *g*_*mid*_ is the middle age of the age group *g* and 4λ represents the total FOI across all four dengue serotypes, consistent with our notation in the previous section. Using a Bayesian framework, the model was fitted to the cross-sectional seroprevalence survey data, which contain age-stratified proportions of the populations who were serologically (IgG) positive.

### Further sensitivity analyses

We also assessed the sensitivity of model estimates based on different data availability:

1. Age stratification. We compared model estimates using different age-binning schemes in Jakarta by comparing the estimates using the original nine age groups and collapsing the data into four age groups matching West Java’s stratification.
2. Temporal data requirement. We assessed minimum data requirements for implementing these models by fitting models to reduced temporal datasets of recent four years and two years of data.

## Results

**Figure 1** illustrates the spatiotemporal patterns and age distribution of reported dengue cases in Jakarta and West Java provinces. The temporal trends (**Figure 1A**) showed distinct patterns between the two provinces. At the province level, Jakarta had higher incidence rates, with peaks in 2019 and 2022, reaching up to 200 cases per 100,000 population. In contrast, West Java maintained generally lower incidence rates, though, at the district level, several districts experienced sporadic high-incidence periods exceeding 300 cases per 100,000 population. The Thousand Island district in Jakarta had the lowest incidence rates among all districts in Jakarta province.

**Figure 1.**
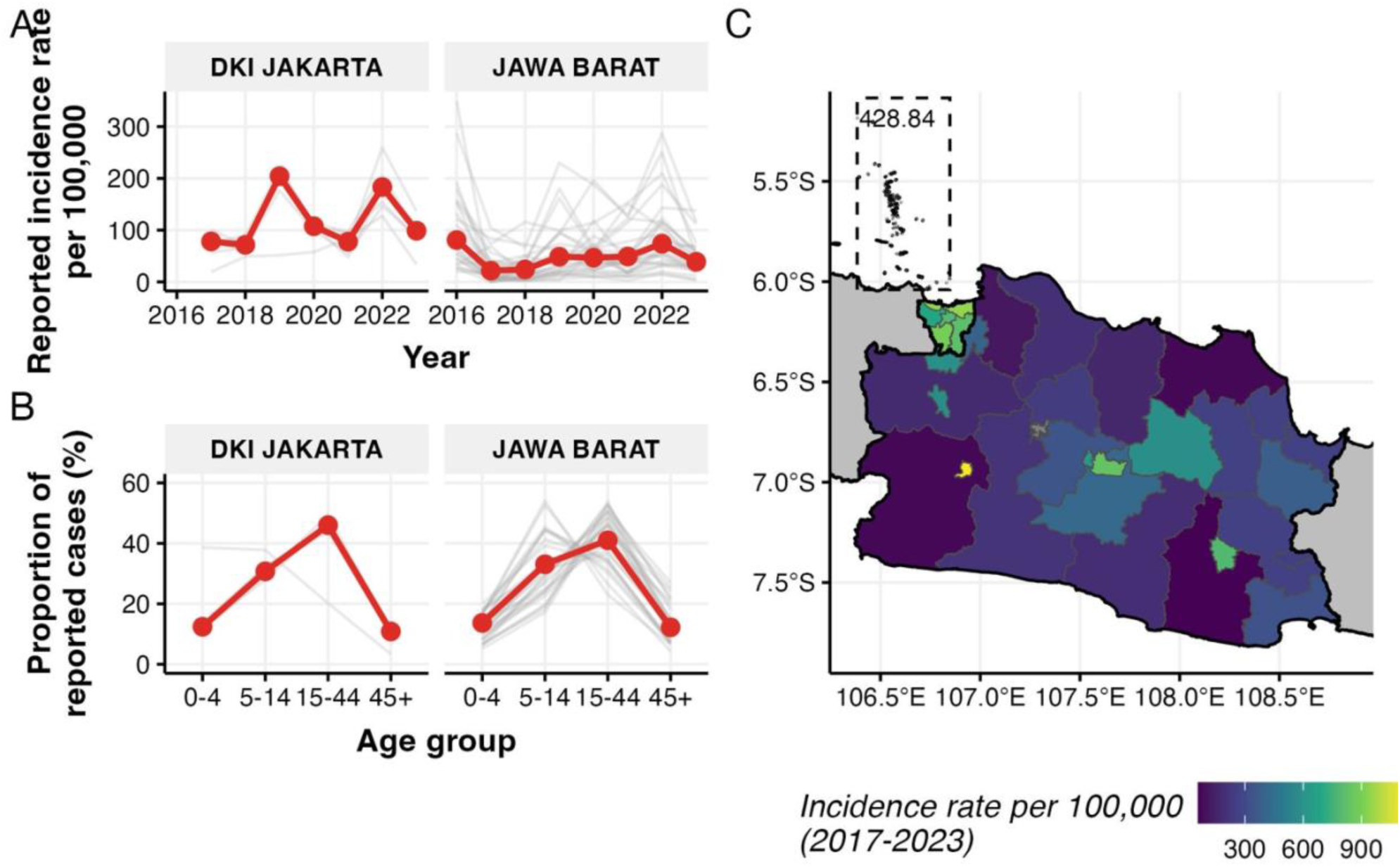
Reported dengue incidence patterns in Jakarta and West Java provinces. (A) Annual reported incidence rates per 100,000 population from 2016-2023. (B) Age distribution of reported cases as a percentage of total cases over the period of observations. Gray lines represent individual districts; red lines highlight one representative district per province. (C) Spatial distribution of overall incidence rates between 2017-2023 at the district level, with the Thousand Island district of Jakarta highlighted in the box with the respective value of incidence rate. Note different scales of incidence rates between temporal trends and spatial distribution. Gray lines denote district boundaries, while black lines denote province boundaries.

The age distribution of cases (**Figure 1B**) showed similar patterns across both provinces, with the highest proportion of cases consistently observed in the 15-44 age group. This age group accounted for approximately 45% of all reported cases in Jakarta and 40% in West Java. The age distributions showed more variation between districts in West Java compared to Jakarta, particularly in the 5-14 and 15-44 age groups. In Jakarta, the Thousand Island district shows different age distribution patterns of cases, with cases reported the highest in the 0-4 and 5-14 age groups, compared to other districts in the province.

The spatial distribution of incidence rates (**Figure 1C**) highlighted substantial heterogeneity across districts, particularly in West Java. Several districts in central areas of West Java reported higher average incidence rates (indicated by lighter colours), while most districts maintained lower rates throughout the study period (darker colours). Jakarta’s districts showed more homogeneous incidence rates compared to West Java, except for the Thousand Island district in the north.

Model performance comparisons using Expected Log Pointwise Predictive Density (ELPD) showed distinct patterns across model structures and provinces (**Figure 2**). In Jakarta (**Figure 2A**), the negative binomial models (Models 2 and 4) showed better predictive performance than their Poisson counterparts (Models 1 and 3). Thousand Island district showed notably better performance (ELPD near -100) under both non-hierarchical specifications regardless of the likelihood functions. Similar patterns were observed in West Java (**Figure 2B**), though with generally higher ELPD values across all models. The negative binomial models again demonstrated better performance, with more compact distributions of ELPD values.

**Figure 2.**
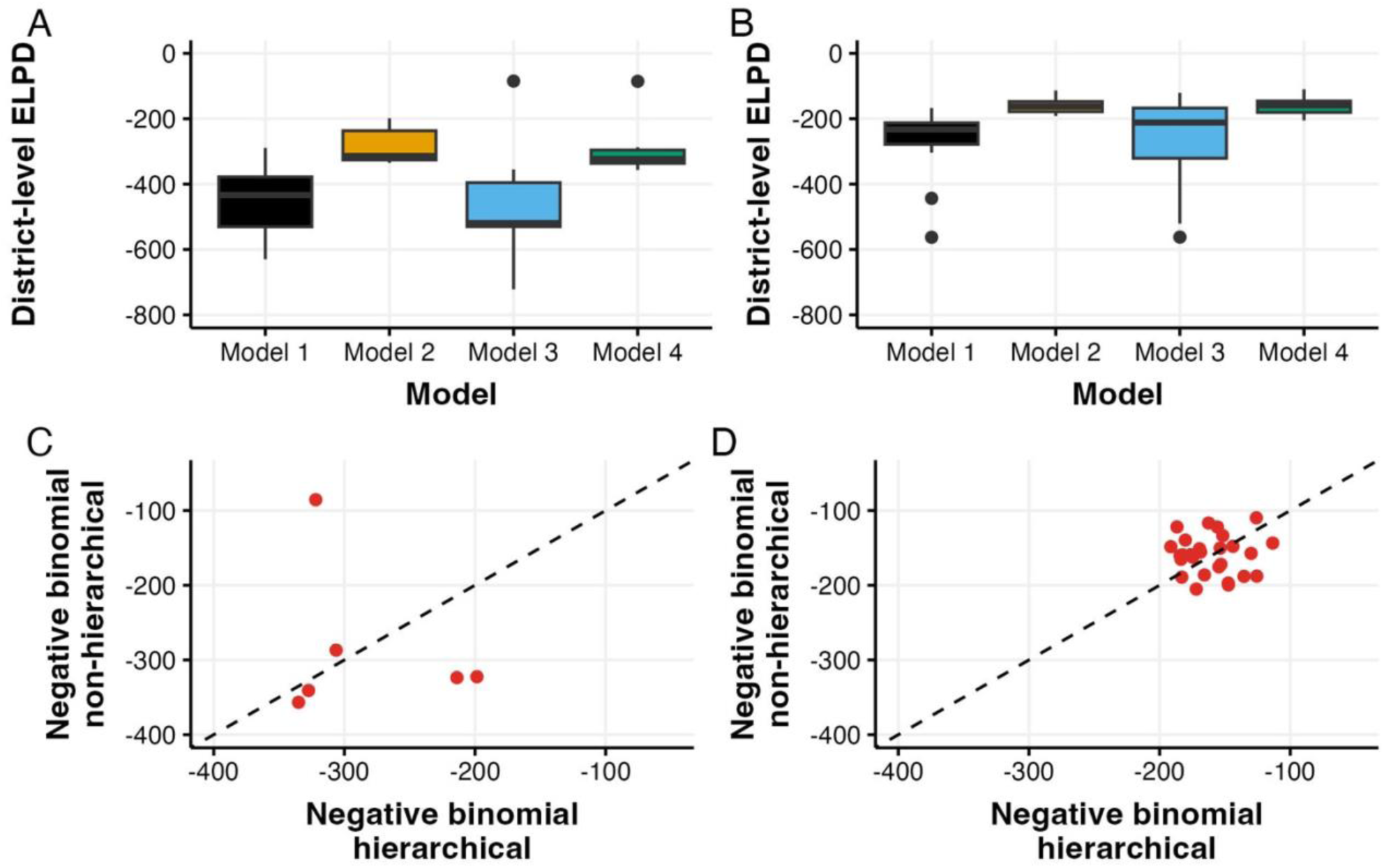
Comparison of model performance using Expected Log Pointwise Predictive Density (ELPD) across four models: Model 1 (Poisson hierarchical), Model 2 (Negative binomial hierarchical), Model 3 (Poisson non-hierarchical), and Model 4 (Negative binomial non-hierarchical). (A) ELPD distributions for Jakarta districts. (B) ELPD distributions for West Java districts. (C, D) Direct comparison of hierarchical versus non-hierarchical negative binomial models for Jakarta (C) and West Java (D), with dashed lines indicating equal performance.

Direct comparison between hierarchical and non-hierarchical negative binomial models (the two best-performing models) showed varying patterns between provinces. In Jakarta (**Figure 2C**), the six districts showed considerable spread around the line of equal performance, suggesting the heterogeneous benefits of having different model structures. In contrast, West Java districts (**Figure 2D**) clustered more tightly around the equality line, with ELPD values predominantly between -150 and -200, indicating more consistent performance between hierarchical and non-hierarchical approaches.

Predicted age-specific seroprevalence patterns from both models closely tracked the 2014 survey data across districts where surveys were conducted (**Figure 3A,B**). In Jakarta, both models captured the steep increase in seroprevalence with age, with estimates largely falling within the credible intervals of the observed data. The age-specific patterns in West Java districts showed greater heterogeneity, ranging from Kota Bandung, with very high seroprevalence starting from 9 years old, to Tasikmalaya, where seroprevalence remained relatively low across ages. The non-hierarchical models produced more district-specific patterns, while hierarchical model estimates showed similar patterns across districts.

**Figure 3.**
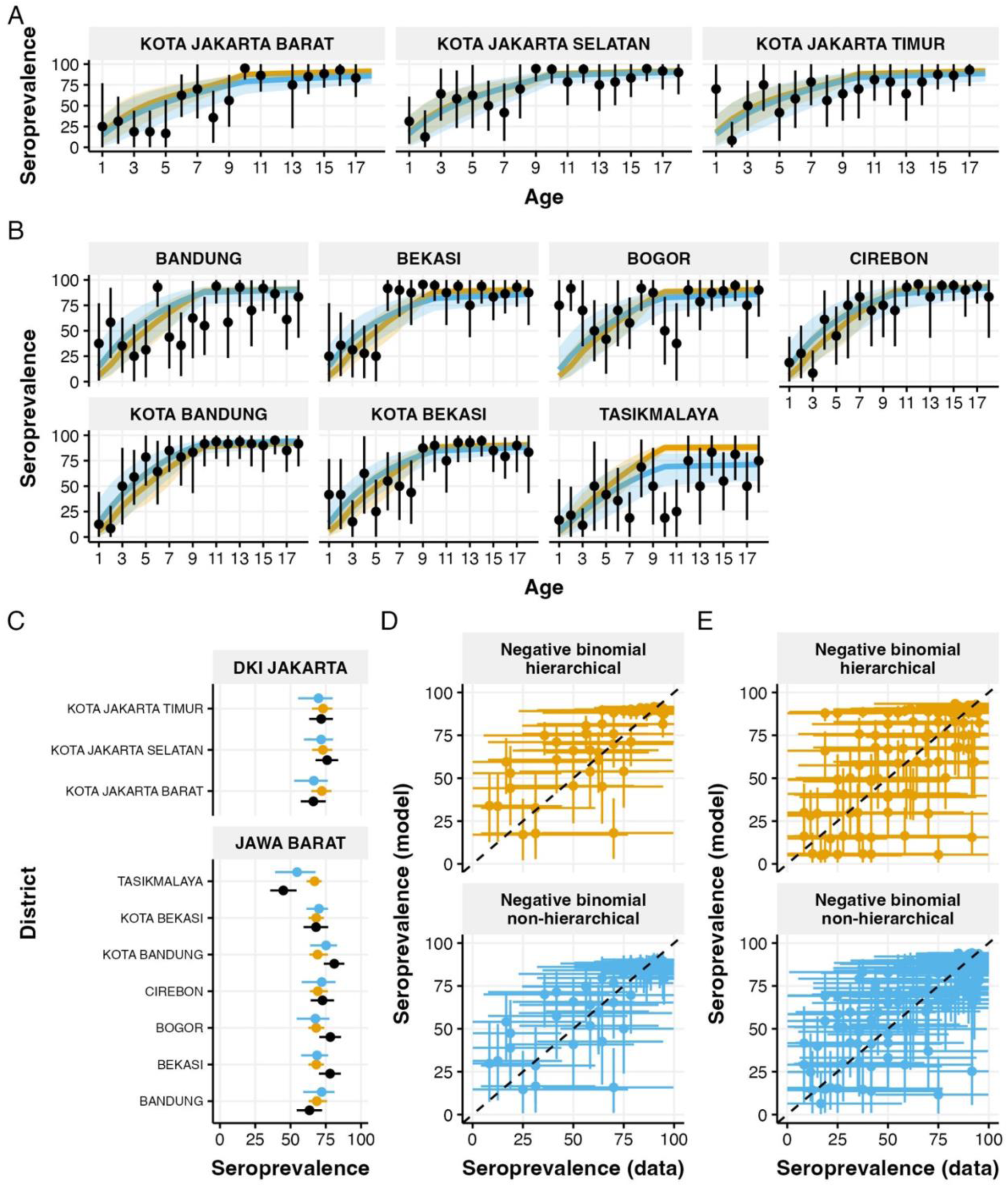
Validation of model estimates against 2014 seroprevalence survey data. (A,B) Age-specific seroprevalence comparisons for Jakarta (A) and West Java (B) districts. Points with error bars show observed seroprevalence data; lines show model estimates (orange: hierarchical negative binomial, blue: non-hierarchical negative binomial) with 95% credible intervals. (C) Overall seroprevalence (ages 1-18 years) comparing model estimates (orange: hierarchical, blue: non-hierarchical) with observed data (black) for each district. (D,E) Model performance assessment showing predicted versus observed seroprevalence values for Jakarta (D) and West Java (E), with dashed lines indicating perfect prediction. All seroprevalences are presented as percentages. Error bars represent 95% credible intervals.

Comparison of overall seroprevalence for ages 1-18 years (**Figure 3C**) showed general agreement between model estimates and survey data across all districts. The hierarchical and non-hierarchical models produced similar estimates in most districts, with the hierarchical model showing narrower credible intervals. In the districts with overall seroprevalence were either the lowest (Kota Jakarta Barat, Tasikmalaya) or the highest (Kota Bandung), the estimates from non-hierarchical models were closer to the observed values. When comparing predicted versus observed seroprevalence values, both models performed similarly in Jakarta (**Figure 3D**) and West Java (**Figure 3E**). The non-hierarchical models produced more scattered predictions, reflecting district-specific estimates, while the hierarchical models generated more clustered predictions, indicating similar estimates for districts within the same province.

**Figures 4A** and **4B** show historical and current estimates of FOI as well as FOI estimates from 2014 seroprevalence survey data for districts where surveys were conducted, with complete results for all districts available in **Figure S1**. Overall, the historical FOI estimates show wide credible intervals, with medians of the historical estimates from non-hierarchical models being closer to estimates from seroprevalence surveys. Furthermore, estimates from non-hierarchical models show district-level heterogeneity, while hierarchical model estimates show similar patterns in historical FOI across all districts within the same province. For current FOI estimates, non-hierarchical model estimates are consistently higher than hierarchical model estimates. This is also highlighted in **Figure 4C**, which contains all districts in both provinces.

**Figure 4.**
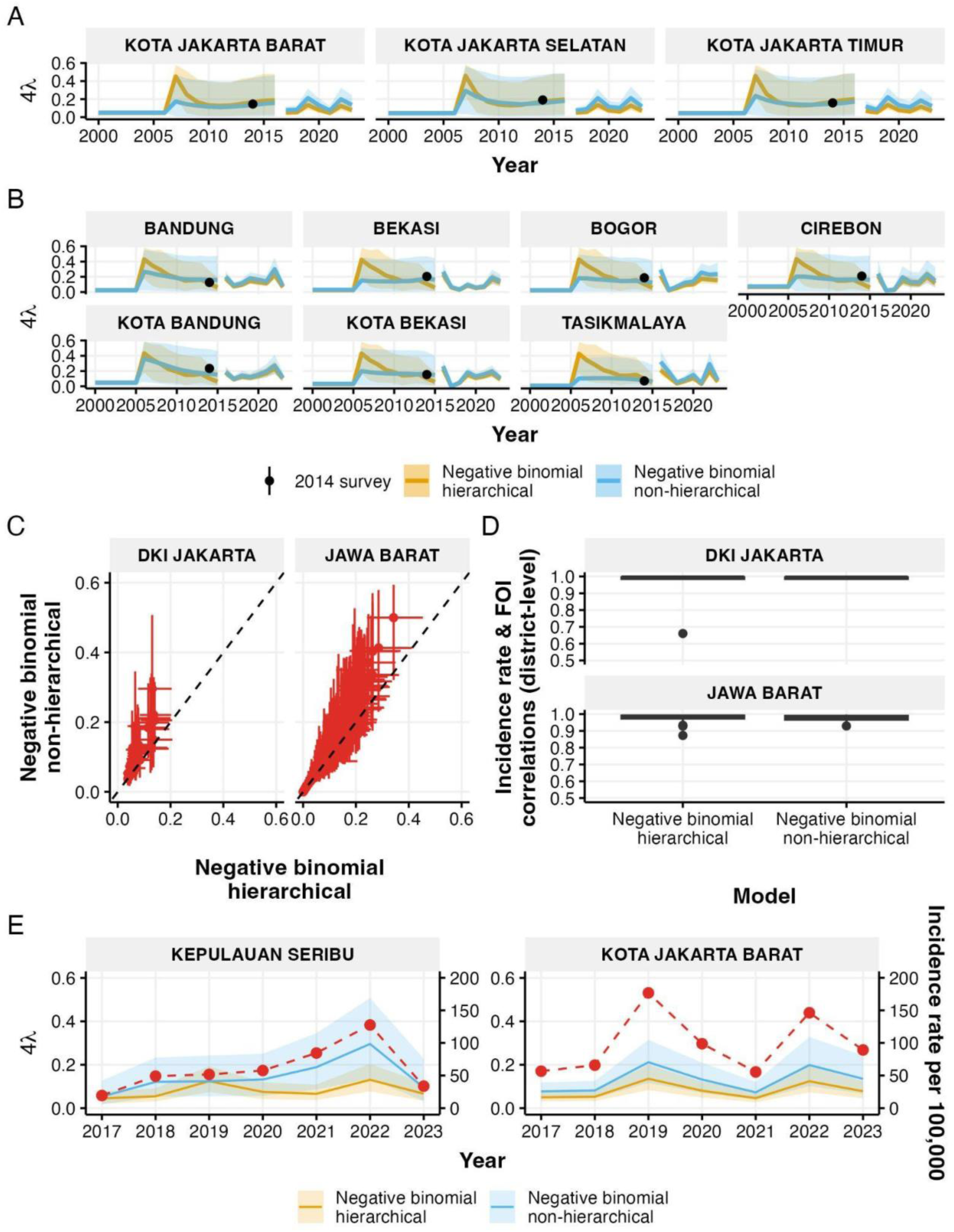
Historical and current force of infection (FOI) estimates in districts with 2014 estimates of FOI from seroprevalence data. (A,B) Historical and current FOI estimates for districts in Jakarta (A) and West Java (B), showing both hierarchical (orange) and non-hierarchical (blue) negative binomial models with 95% credible intervals. Black points indicate FOI estimates from 2014 seroprevalence surveys. (C) Comparison of FOI estimates between hierarchical and non-hierarchical models for all districts in Jakarta (left) and West Java (right), with error bars showing 95% credible intervals. (D) District-level correlations between FOI estimates and reported incidence rates for both models. The detailed district-level patterns of these two variables are available in **Figure S2,3**. (E) Detailed comparison of FOI estimates and incidence rates (red dashed line) in two contrasting districts: Thousand Island district showing low correlation (Jakarta’s outlier in (D)) and West Jakarta City showing high correlation between FOI and incidence rate.

District-level correlations between FOI estimates and reported incidence rates showed high consistency across most districts (**Figure 4D**). However, in Jakarta, the Thousand Island district showed a notably lower correlation, particularly for the hierarchical model. This difference is illustrated in **Figure 4E**, where the pattern of hierarchical model estimates of FOI for the Thousand Island district showed less alignment with local incidence patterns than the non-hierarchical model. In contrast, in West Jakarta City, there is a strong correlation between FOI estimates and incidence rates in both models, with FOI patterns closely tracking temporal changes in reported incidence.

**Figure 5A** shows that the reporting parameter (ρ) estimates were generally higher under the hierarchical model than the non-hierarchical model, particularly for districts in Jakarta. The relative reporting ratio (γ) showed more alignment between models, though with wider uncertainties in the non-hierarchical model estimates, with a district showing a median estimate above 0.75 (the Thousand Island district in Jakarta). In contrast, hierarchical model estimates remained mostly below 0.5 (**Figure 5B**).

**Figure 5.**
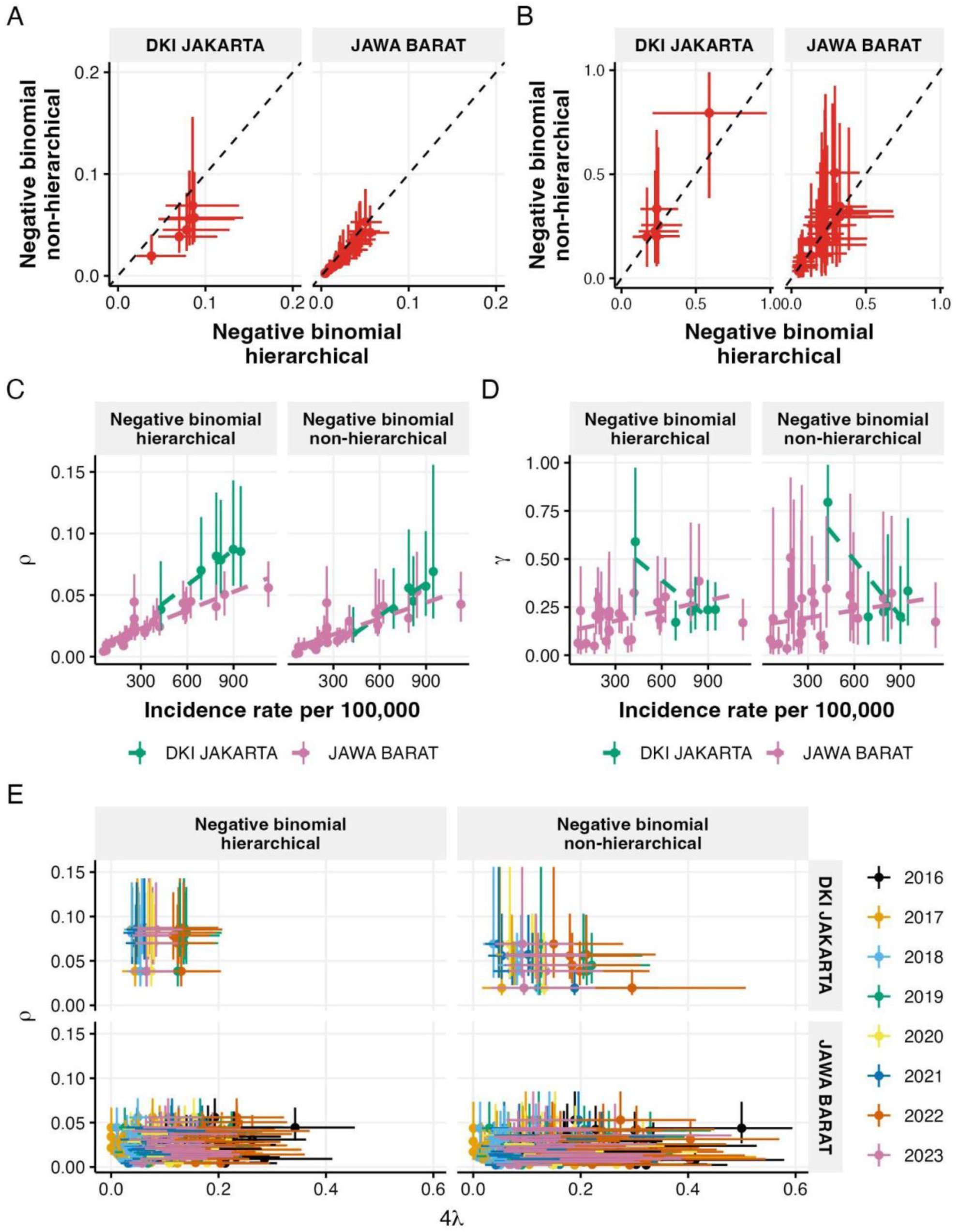
Parameter estimates and their relationships. (A,B) Comparison between hierarchical and non-hierarchical model estimates of reporting parameter, ρ, (A) and relative reporting ratio, γ, (B) for districts in Jakarta and West Java, with error bars showing 95% credible intervals. (C,D) Relationships between incidence rates and parameter estimates: ρ (C) and γ (D), shown separately for hierarchical and non-hierarchical models. (E) Relationship between ρ and FOI across years, comparing hierarchical (left) and non-hierarchical (right) models for both provinces.

When examining the relationship between parameters and incidence rates (**Figure 5C,D**), both models showed positive correlations between ρ and incidence rates, with Jakarta districts displaying generally higher reporting parameters than West Java districts. However, the non-hierarchical model showed more similar ρ and incidence rate relationships between provinces than the hierarchical model. For γ, there was almost no correlation with incidence rates in either province or model, especially when excluding the outlier estimates from the Thousand Island district in Jakarta.

The relationship between ρ and FOI showed no correlations in most cases (**Figure 5E**, with detailed posterior sample-level relationships in **Figure S4,5**), except for a slight negative correlation in Jakarta districts under the non-hierarchical model. Despite having generally lower FOI values with narrower ranges, Jakarta districts consistently showed higher ρ estimates than West Java districts.

To assess the impact of age stratification on parameter estimation, we compared model performance using Jakarta’s detailed nine age groups versus the condensed four age groups used in other provinces (**Figure 6**). Historical FOI estimates showed similar temporal patterns between both age-grouping schemes, maintaining consistency with 2014 seroprevalence data points (**Figure 6A**, complete results for all districts available in **Figure S6**). Both age-grouping schemes produced estimates consistent with the 2014 seroprevalence data points, although median estimates from non-hierarchical models show more consistency between different age groupings. The hierarchical model showed systematic differences between age-grouping schemes, with four age groups consistently producing higher seroprevalence and reporting rate (ρ) estimates but lower relative reporting ratios (γ) compared to nine age groups (**Figures 6B-E**). These differences, while falling within uncertainty bounds, suggest the hierarchical structure may be more sensitive to age stratification choices. In contrast, the non-hierarchical model showed more consistent median estimates between age-grouping schemes across all parameters, though with wider credible intervals for the four age-group structure.

**Figure 6.**
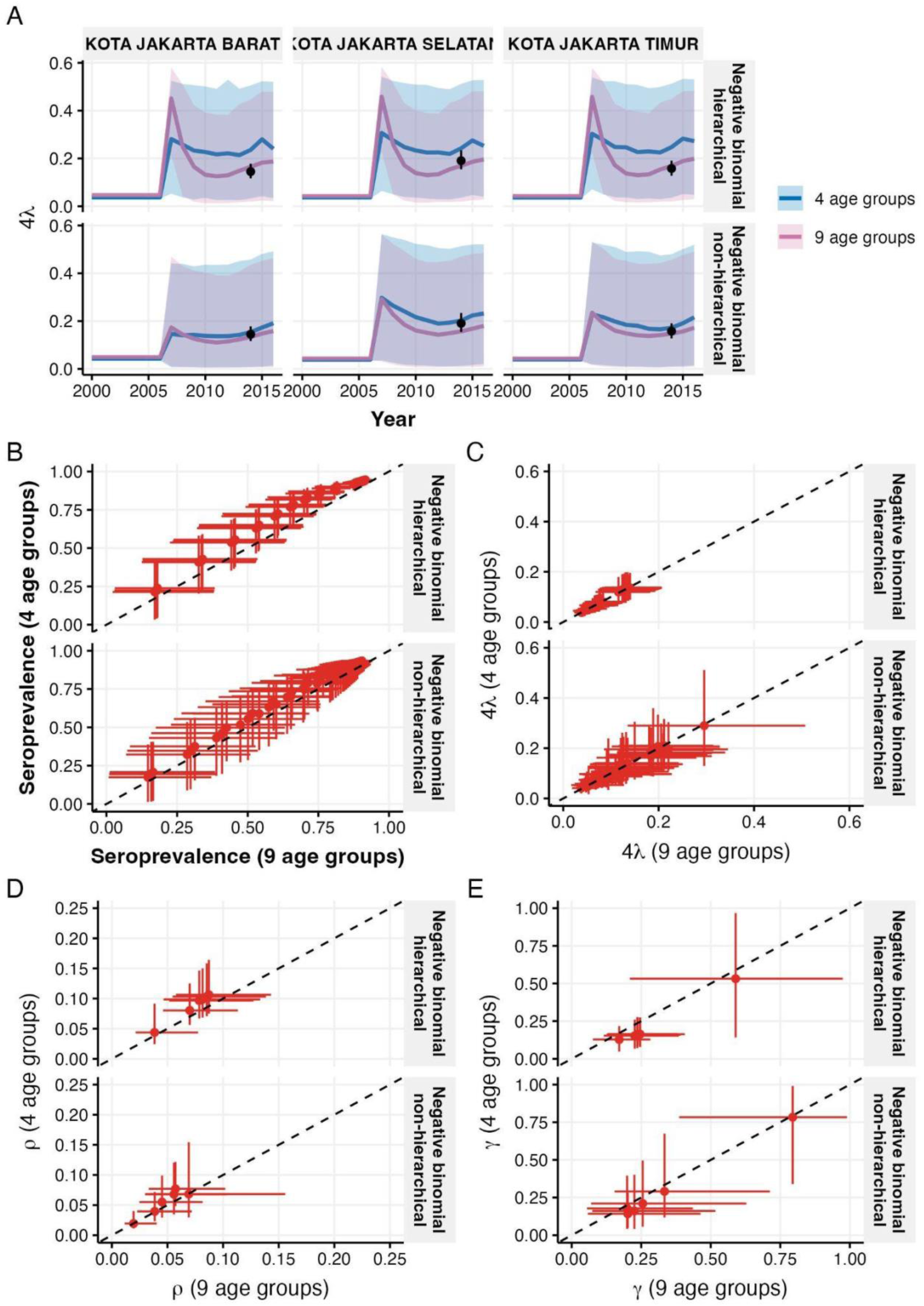
Sensitivity analysis comparing model estimates using four versus nine age groups for Jakarta districts. (A) Historical FOI estimates from hierarchical (top) and non-hierarchical (bottom) models, with black points showing 2014 seroprevalence-derived estimates. (B,C) Comparison of age-specific seroprevalence (B) and FOI estimates (C) between models using different age groupings. (D,E) Comparison of reporting parameter ρ (D) and relative reporting ratio γ (E) estimates between models using different age groupings. Dashed lines indicate perfect agreement; error bars show 95% credible intervals.

To assess the practical implications of model choice for intervention targeting, we mapped the modelled probability of districts exceeding the WHO-defined high transmission threshold (seroprevalence >60% in 9-year-olds [27]) for both 2017 and 2023 (**Figure 7**). In 2017, both hierarchical (**Figure 7A**) and non-hierarchical (**Figure 7B**) models showed consistently high probabilities (>95%) across nearly all districts in both provinces, indicating widespread high transmission conditions after a huge dengue outbreak nationwide in 2016. However, by 2023, both models diverged in their predictions. The non-hierarchical model (**Figure 7C**) maintained high transmission probabilities across most districts, with two districts (Kota Jakarta Utara and Karawang) showing 90% probabilities (84% and 89%, respectively). In contrast, the hierarchical model (**Figure 7D**) showed much greater spatial heterogeneity, with eight districts displaying <90% probabilities (the lowest, Karawang, has 75% probability). A direct comparison showed only 75% agreement between the two approaches when classifying districts as high-transmission areas using a 90% probability threshold in 2023.

**Figure 7.**
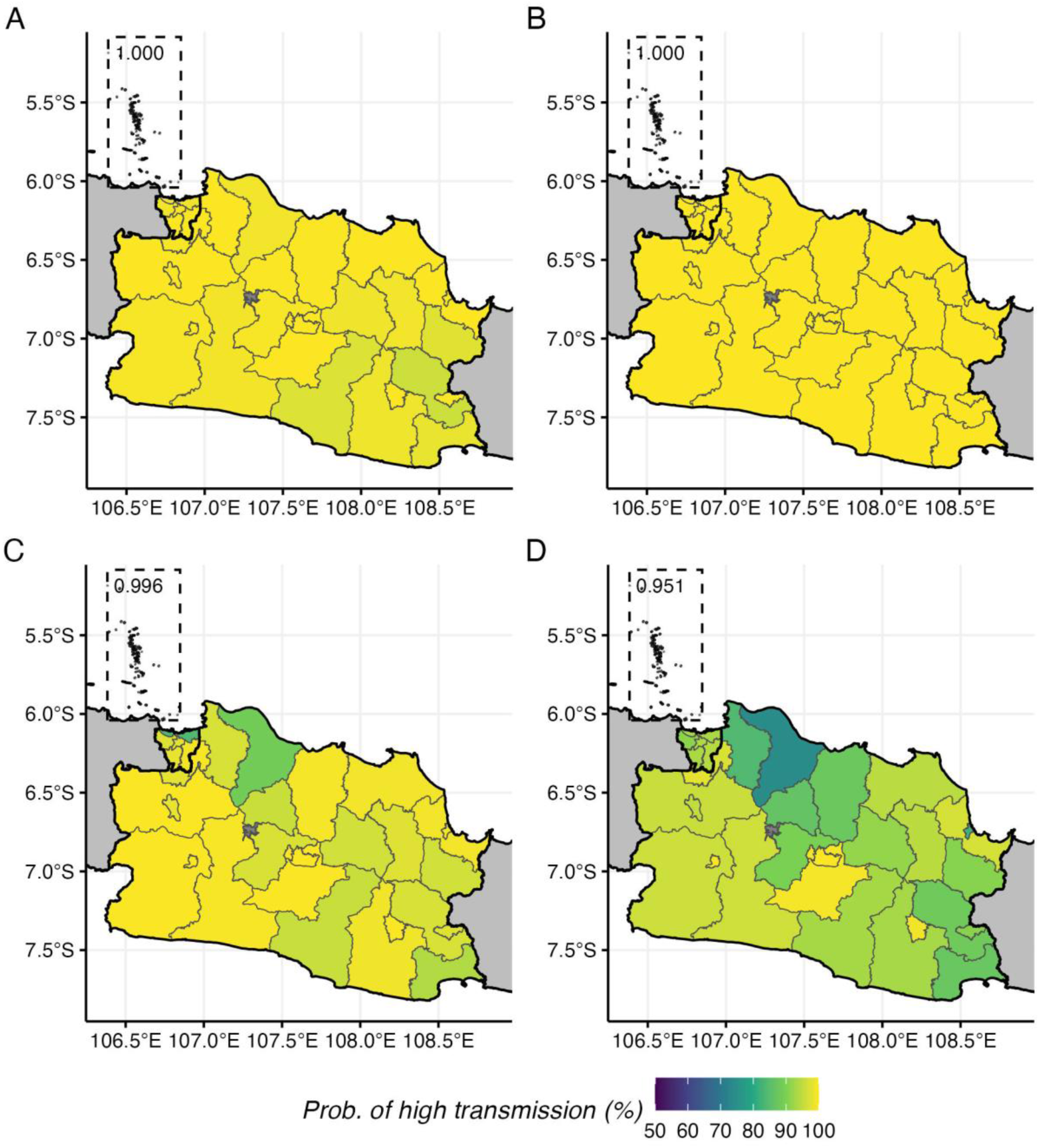
Map of modelled probability of having high dengue transmission at the district level. (A) In 2017 based on non-hierarchical model. (B) In 2017 based on hierarchical model. (C) In 2023 based on non-hierarchical model. (D) In 2023 based on hierarchical model.

## Discussions

In this study, we developed and compared hierarchical and non-hierarchical catalytic models to estimate dengue FOI using age-stratified surveillance data from districts in Jakarta and West Java provinces. Both modelling approaches produced historical FOI estimates consistent with 2014 seroprevalence data, though non-hierarchical models showed closer alignment with seroprevalence-derived estimates in individual districts. The hierarchical framework produced narrower credible intervals and more stable estimates across districts within provinces. However, this came at the cost of potentially missing district-specific transmission patterns, as exemplified by the Thousand Islands district where local incidence patterns differed substantially from other Jakarta districts. Model comparison using ELPD showed that negative binomial models performed better than Poisson models in both hierarchical and non-hierarchical frameworks, indicating the importance of accounting for overdispersion in the data. Our analysis showed distinct patterns in reporting parameters between provinces, with districts in Jakarta showing generally higher reporting rates despite lower FOI estimates compared to districts in West Java, reflecting differences in surveillance system capabilities and healthcare access between these provinces.

This study firstly highlights the importance of considering overdispersion in the reported incidence data by using negative binomial distribution as the likelihood function for model fitting. The negative binomial models greatly outperform their Poisson counterparts regardless of whether hierarchical structure was considered. This is not surprising as disease surveillance data often show overdispersion due to various factors including heterogeneity in individual susceptibility and transmissibility, clustering of cases, and variability in healthcare-seeking behaviour and reporting practices [28–30], and previous modelling studies have already used negative binomial likelihood to handle overdispersion [16–18]. While our analysis demonstrated better performance of negative binomial likelihood in handling overdispersion, future studies could explore alternative approaches such as generalised Poisson distribution [31] or quasi-Poisson likelihood [32]. These approaches may offer different trade-offs between model complexity and ability to capture overdispersion in disease surveillance data.

When considering using hierarchical and non-hierarchical approaches, we need to consider potential trade-offs linked with variations in local epidemiology of dengue within regions of interest. Hierarchical models provide better precision in estimates and may help districts with low count of data within the same region. However, when an outlier district such as the Thousand Island district in Jakarta exists, we may miss district-specific patterns as the partial pooling of the hierarchical structure pulls estimates toward the majority patterns from other districts within the region [33]. Additionally, this partial pooling effect consistently produces lower FOI estimates in hierarchical models compared to non-hierarchical models, with the difference appearing to be compensated by correspondingly higher reporting rate estimates, suggesting an identifiability trade-off between these parameters when fitting to the same surveillance data.

Understanding local epidemiology and considering whether districts within a region share similarities is crucial when implementing hierarchical modelling, as it is not a given that all districts within the region of interest are similar in their transmission patterns. Our study also highlights the importance of validating model estimates against seroprevalence data or other datasets outside surveillance data. We showed how hierarchical models produced similar historical patterns across districts within the same provinces that may not capture district-specific seroprevalence patterns as well as the non-hierarchical models. This is an important aspect to consider as historical transmission patterns affect how immunity is accumulated in the population and subsequently influence the outbreak patterns in the future [34], even though for dengue, it may not be as straightforward due to the serotypes dynamics [35,36]. While our validation relied on 2014 seroprevalence data collected before our study period, ideally, future implementations would benefit from concurrent seroprevalence surveys conducted within the period of surveillance data collection to provide more direct validation of contemporary FOI estimates.

Understanding parameter estimates’ drivers and their interpretations is crucial for translating model outputs into public health decisions. We found several key relationships between model parameters and surveillance data. First, FOI estimates showed a strong correlation with reported incidence rates, which is expected given that in our model, FOI directly influences infection occurrences while the constant reporting parameter adjusts the proportion of these infections that are reported. The reporting parameter (ρ) also strongly correlated with overall incidence rates across the study period, with the highest reporting rates observed in districts with ’city’ status or the more urbanised areas within each province. This pattern aligns with the expected better healthcare access and surveillance systems in urban areas compared to non-urban districts [37]. The relative reporting ratio (γ) showed no correlations with incidence rates. However, with its distinct epidemiology, the Thousand Island district showed notably higher γ estimates. The relatively low level of estimates of γ in most districts suggests long-endemic dengue areas where secondary infections dominate reported cases [25]. In the Thousand Island district, both primary and secondary infections were estimated to contribute substantially to reported cases, suggesting a different epidemiological state than other districts, an observation supported by its uniquely high proportion of infant cases.

Regarding parameter identifiability, particularly the potential negative correlation between FOI and reporting parameters, our analysis showed largely independent relationships in posterior samples, except for a slight negative correlation in Jakarta’s non-hierarchical model. Importantly, validating historical seroprevalence estimates against observed data increases confidence in the reliability of both FOI and reporting parameter estimates.

Our analysis shows that selecting between hierarchical and non-hierarchical models for analysing age-stratified dengue case data requires careful consideration of both local epidemiology and data quality. In its basic form (as implemented in this work), the hierarchical model is most suitable for regions where local epidemiology between districts is relatively homogeneous. However, the presence of even a single district with distinct dengue epidemiology and transmission dynamics may result in estimates that fail to reflect its unique local patterns. The hierarchical approach also shows sensitivity to data resolution, with different age-stratification schemes producing slightly different estimates, potentially leading to implementation inconsistencies depending on data availability. Nevertheless, its key advantage lies in producing estimates with narrower uncertainty bounds through partial pooling of parameters across districts. Non-hierarchical models demonstrate more consistent parameter estimates across different levels of data detail and better capture district-specific epidemiological patterns. However, they produce wider uncertainty bounds compared to hierarchical models, particularly with less detailed data. While four age groups can provide useful parameter estimates, more detailed age stratification reduces estimation uncertainty. More temporal data length also improves estimates’ reliability. Further analysis shows that hierarchical models suffer from inconsistency in estimates when different lengths of data were used (**Figure S7-9**). Regardless of the chosen approach and data availability, model implementation should incorporate validation against external datasets, comparison with other epidemiological indicators, and careful assessment of unusual patterns in model estimates to ensure reliability.

Our model assumes full susceptibility to all four dengue serotypes in the youngest age group (0-4 years old), which may not accurately reflect the epidemiological situation for infants under 1 year of age. Maternal dengue antibodies can provide temporary protection against infection but may also enhance disease severity through antibody-dependent enhancement, potentially affecting both infection patterns and reporting rates in this population. The inability to separate infant data from our aggregated 0-4 years old age group represents a limitation of our current analysis.

We demonstrate a practical application of our modelling approach for intervention planning by simulating actionable metrics from catalytic models using posterior samples of fitted models. The latest WHO-recommended dengue vaccine is indicated for use in high-transmission areas, defined as those with dengue seroprevalence exceeding 60% in 9-year-olds [27]. While getting information about population immunity traditionally requires resource-intensive seroprevalence surveys, the catalytic modelling framework, such as the one presented here and other studies [15–19], enables estimating population immunity profiles and age-specific seroprevalence using routine surveillance data. The 25% disagreement in district classifications between non-hierarchical and hierarchical modelling approaches highlights the practical impact of methodological choices on intervention-targeting decisions. This methodological uncertainty becomes particularly important when the decisions involve substantial financial commitments or when targeting errors could result in missed opportunities to prevent transmissions or cases.

While accounting for the caveats in data quality and model selection discussed above, these probability of high-transmission area estimates can provide valuable additional evidence for strategic intervention planning, particularly in resource-limited settings where comprehensive seroprevalence surveys may not be feasible. Additionally, the FOI estimates could serve as reference points for evaluating the impact of ongoing Wolbachia releases in five Indonesian districts [5] by enabling a comparison of transmission intensity before and after intervention.

Beyond intervention planning and evaluation, our FOI estimates provide important information for calculating the subnational dengue infection burden in Indonesia, including undetected and unreported cases. By comparing reported cases with the estimated total infections derived from FOI that accounts for reporting parameter (ρ), health authorities can quantify the hidden burden of dengue across different regions. This approach helps reveal the true magnitude of dengue transmission beyond what surveillance systems capture, offering a more accurate basis for resource allocations and surveillance strengthening programmes. To our knowledge, this study represents the first comprehensive application of catalytic modelling approaches to estimate subnational dengue FOI using routine surveillance data. Developing a pipeline for a nationwide implementation of FOI estimation framework using routine surveillance data with annual updates would strengthen existing monitoring mechanisms specified in Indonesia’s National Strategic Plan for dengue control [4].

While our study incorporates time-varying FOI estimates, these estimates are primarily reliable for years with available age-stratified case data. Although we estimated historical FOI (both recent and longer-term estimates from before data collection began), these historical estimates carried substantial uncertainty and could not capture the detailed transmission dynamics of those earlier periods. Similarly, the available seroprevalence survey data from 2014 had insufficient age coverage and sample sizes to reliably estimate time-varying FOI patterns. These limitations highlight the need for methodological advances and improved data collection strategies to better estimate historical time-varying FOI of dengue.

Developing robust estimates of historical time-varying FOI would offer significant advantages for understanding dengue epidemiology while also reflecting the complex nature of dengue transmission dynamics in the population [38]. Such estimates would enable an analysis of relationships between FOI and historical changes in climate, environmental conditions, urbanisation levels, and socioeconomic factors. This understanding could improve our ability to predict future dengue transmission dynamics and intensity in response to projected changes in these variables, ultimately strengthening the evidence for intervention planning and policy decisions.

Finally, we propose several directions for future work to improve FOI estimation using catalytic models, particularly within the hierarchical modelling framework and Indonesian context. First, incorporating covariates directly related to dengue transmission, such as urbanicity and population density, could better inform heterogeneity between districts within regions or provinces. Second, integrating diverse data sources would strengthen our approach. For example, long-term temporal dengue case data (without age stratification) could be combined with newer age-stratified surveillance data to better inform historical time-varying FOI patterns, potentially improving current estimates. Third, future implementations would benefit from access to more detailed age-stratified surveillance data, particularly with separate reporting for infants under 1 year, to better account for the complex immunological dynamics in this vulnerable population. Fourth, exploring alternative prior distribution assumptions for hierarchical parameter estimation of FOI (such as Gamma distribution) could potentially improve the framework’s ability to accommodate outlier districts while maintaining the benefits of partial pooling. Our review of publicly available dengue-related data in Indonesia identified multiple valuable but fragmented dengue-related data, including seroprevalence surveys, case serotype information, and longitudinal surveillance data. Future development of catalytic modelling approaches should focus on frameworks that coherently integrate these diverse data types to provide more robust estimates of FOI (with emphasis on time-varying and historical FOI) and epidemiological parameters, and better inform policy decisions.

## Acknowledgements

We thank the Arbovirus Working Group at the Ministry of Health of Indonesia for providing access to the surveillance data and their technical support.

## Funding

BAD is supported by funding from the Program for Research in Epidemic Preparedness and Response (PREPARE) from the Ministry of Health, Singapore (A-8000642-01-00 PREPARE S2-2024-002). IRFE is supported by the Wellcome Africa Asia Program Vietnam (106680/Z/I4/Z) and the Strategic Partnership for Prevention, Surveillance and Response to Infectious Diseases across the Indo-Pacific Region (SPARKLE). HEC is supported by NUS Start up Grant awarded to her. SM acknowledges support from the National Research Foundation, Singapore, under its NRF FELLOWSHIP (NRF-NRFF15 - 2023 - 0010) awarded to him.

## Ethics

This work did not require ethical approval from a human subject or animal welfare committee.

## Data accessibility

The codes to perform analyses in this article and data for Jakarta province are available at: Zenodo https://doi.org/10.5281/zenodo.17239255 and GitHub https://github.com/mlgh-sg/catalytic_dengue_java. The routine surveillance data for Jakarta are also freely accessible from: https://surveilans-dinkes.jakarta.go.id/

## Declaration of AI use

We have not used AI-assisted technologies in creating this article.

## Authors’ contributions

BAD: conceptualization, methodology, formal analysis, writing—original draft; IRFE: writing— review & editing; FSMS: data curation, writing—review & editing; AS: writing—review & editing; AH: data curation, writing—review & editing; MK: writing—review & editing; DA: data curation, writing—review & editing; DG: writing—review & editing; H: writing—review & editing; AKN: writing—review & editing; EP: writing—review & editing; IS: writing—review & editing; AS: writing—review & editing; BT: writing—review & editing; HEC: writing—review & editing; SM: conceptualization, methodology, funding acquisition, writing—review & editing.

## Conflict of interest declaration

The authors declare no competing interests.

## Disclaimer

The views in this article are those of the authors and do not necessarily represent the views, decisions, or policies of the institutions with which the authors are affiliated.

## Notes

### Competing Interest Statement

The authors have declared no competing interest.

### Summary of Updates

Moved seroprevalence estimates to main texts

