## Supplementary information for "Estimating dengue force of infection from age-stratified surveillance data in Java, Indonesia"

This is a supplementary information document for a manuscript titled: "***Estimating dengue force of infection from age-stratified surveillance data in Java, Indonesia***"

Bimandra A Djaafara[1,2*], Iqbal RF Elyazar[2,3], Asik Surya[4], Fadjar SM Silalahi[4], Agus Handito[4], Desfalina Aryani[4], Mushtofa Kamal[5], Dyana Gunawan[4], Hipokrates[4], Anzala Khoirun Nisa[4], Edi Prianto[4], Iriani Samad[4], Agus Sugiarto[4], Burhannudin Thohir[4], Hannah E Clapham[1], Swapnil Mishra[1,6,7,8*]

[1]Saw Swee Hock School of Public Health, National University of Singapore and National University Health System, Singapore

[2]Oxford University Clinical Research Unit Indonesia, Faculty of Medicine, University of Indonesia, Indonesia

[3]Monash University Indonesia, Tangerang, Banten, Indonesia

[4]Arbovirus Working Group, Ministry of Health of the Republic of Indonesia, Indonesia

[5]World Health Organization, Country Office for Indonesia, Jakarta, Indonesia

[6]Institute of Data Science, National University of Singapore, Singapore

[7]Department of Statistics and Data Science, National University of Singapore, Singapore

[8]Research Office, National Centre for Infectious Diseases, Singapore

### Supplementary Information

#### Priors selections for the hierarchical and non-hierarchical models

For the hierarchical framework, the model fitting processes were done independently in each province. The province-level priors and district-level priors for $\lambda_{d,H}$, $\lambda_{d,t}$, $\rho$, $\gamma$, and $\phi$ are defined as follows:

$$\lambda_{constant,prov,H} \sim Normal^{+}(0{,}0.05)$$

$$\kappa_{\lambda_{constant,H}} \sim Normal^{+}(0{,}1)$$

$$\lambda_{constant,d,H} \sim Normal^{+}(\lambda_{constant,prov,H}, \kappa_{\lambda_{constant,H}})$$

$$\lambda_{recent,prov,H} \sim Normal^{+}(0{,}0.05)$$

$$\kappa_{\lambda_{recent,H}} \sim Normal^{+}(0{,}1)$$

$$\lambda_{recent,d,H} \sim Normal^{+}(\lambda_{recent,prov,H}, \kappa_{\lambda_{recent,H}})$$

$$\lambda_{prov,t} \sim Normal^{+}(0{,}0.05)$$

$$\kappa_{\lambda_t} \sim Normal^{+}(0{,}1)$$

$$\lambda_{d,t} \sim Normal^{+}(\lambda_{prov,t}, \kappa_{\lambda_t})$$

$$\rho_{prov} \sim Normal^{+}(0{,}0.15)$$

$$\kappa_{\rho} \sim Normal^{+}(0{,}1)$$

$$\rho_{d} \sim Normal^{+}(\rho_{prov}, \kappa_{\rho})$$

$$\gamma_{prov} \sim Normal^{+}(0{,}1)$$

$$\kappa_{\gamma} \sim Normal^{+}(0{,}1)$$

$$\gamma_{d} \sim Normal^{+}(\gamma_{prov}, \kappa_{\gamma})$$

$$\phi_{prov} \sim InverseGamma(0.4{,}0.3)$$

$$\kappa_{\phi} \sim Normal^{+}(0{,}1)$$

$$\phi_{d} \sim Normal^{+}(\phi_{prov}, \kappa_{\phi})$$

For the non-hierarchical framework, the model fitting processes were done independently in each district. The district-level priors for $\lambda_{d,H}$, $\lambda_{d,t}$, $\rho$, $\gamma$, and $\phi$ are defined as follows:

$$\lambda_{constant,d,H} \sim Normal^{+}(0,0.05)$$

$$\lambda_{recent,d,H} \sim Normal^{+}(0,0.05)$$

$$\lambda_{d,t} \sim Normal^{+}(0,0.05)$$

$$\rho_{d} \sim Normal^{+}(0,0.15)$$

$$\gamma_{d} \sim Normal^{+}(0,1)$$

$$\phi_{d} \sim InverseGamma(0.4,0.3)$$

#### Supplementary figures

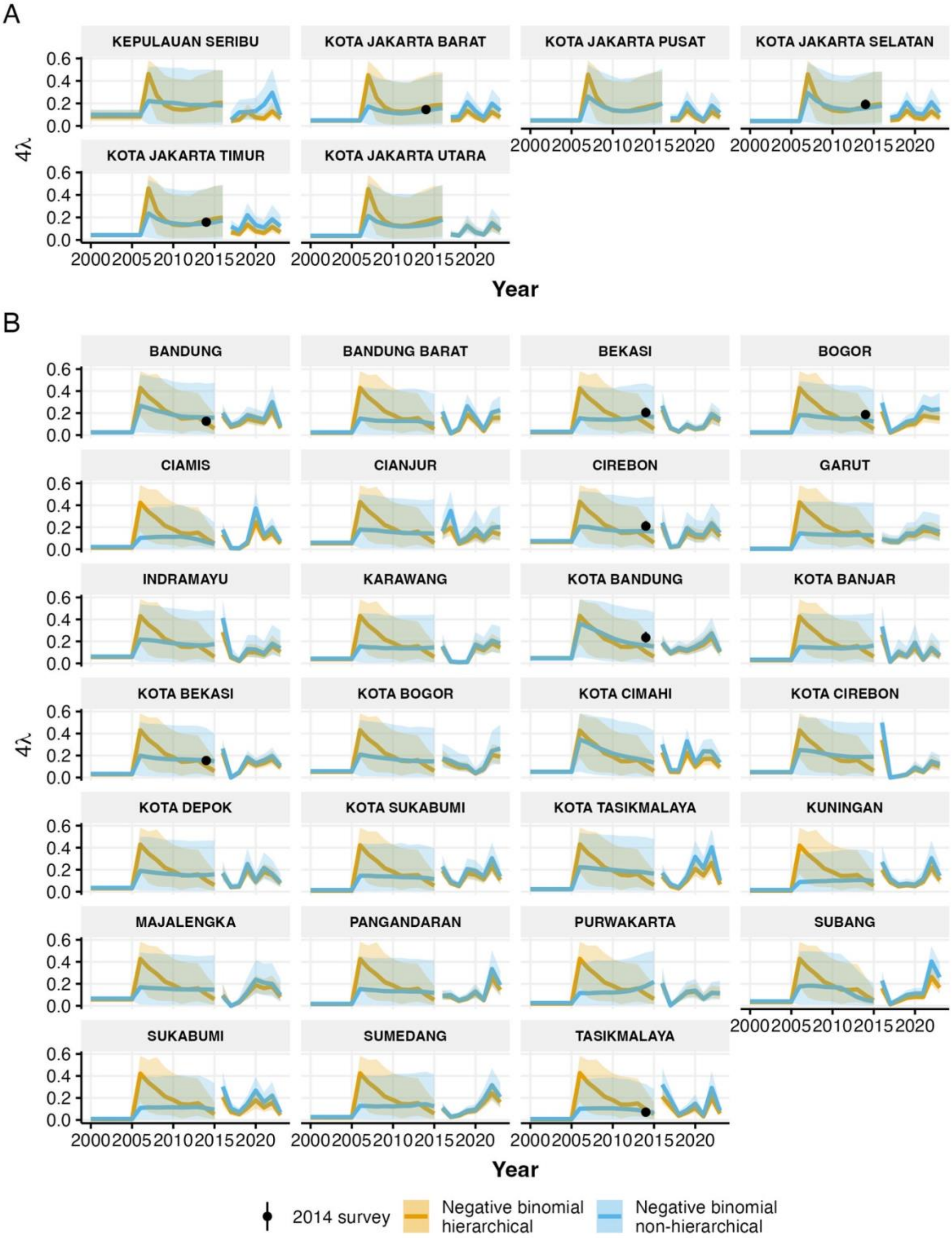

**Figure S1.** Historical and current force of infection (FOI) estimates in districts with 2014 estimates of FOI from seroprevalence data. (A,B) Historical and current FOI estimates for districts in Jakarta (A) and West Java (B), showing both hierarchical (orange) and non-hierarchical (blue) negative binomial models with 95% credible intervals. Black points indicate FOI estimates from 2014 seroprevalence surveys.

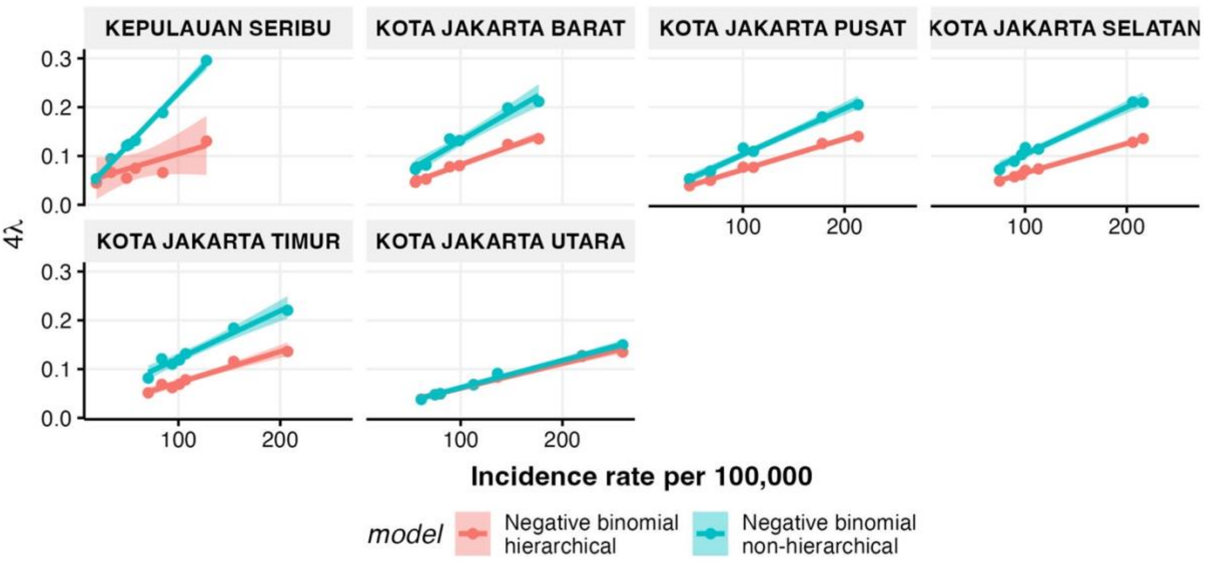


**Figure S2.** Scatter plots of district-level relationship between reported incidence rate and FOI estimates for both hierarchical and non-hierarchical models in Jakarta province. Coloured lines denote the estimated linear regression lines between the two variables from each model estimates.

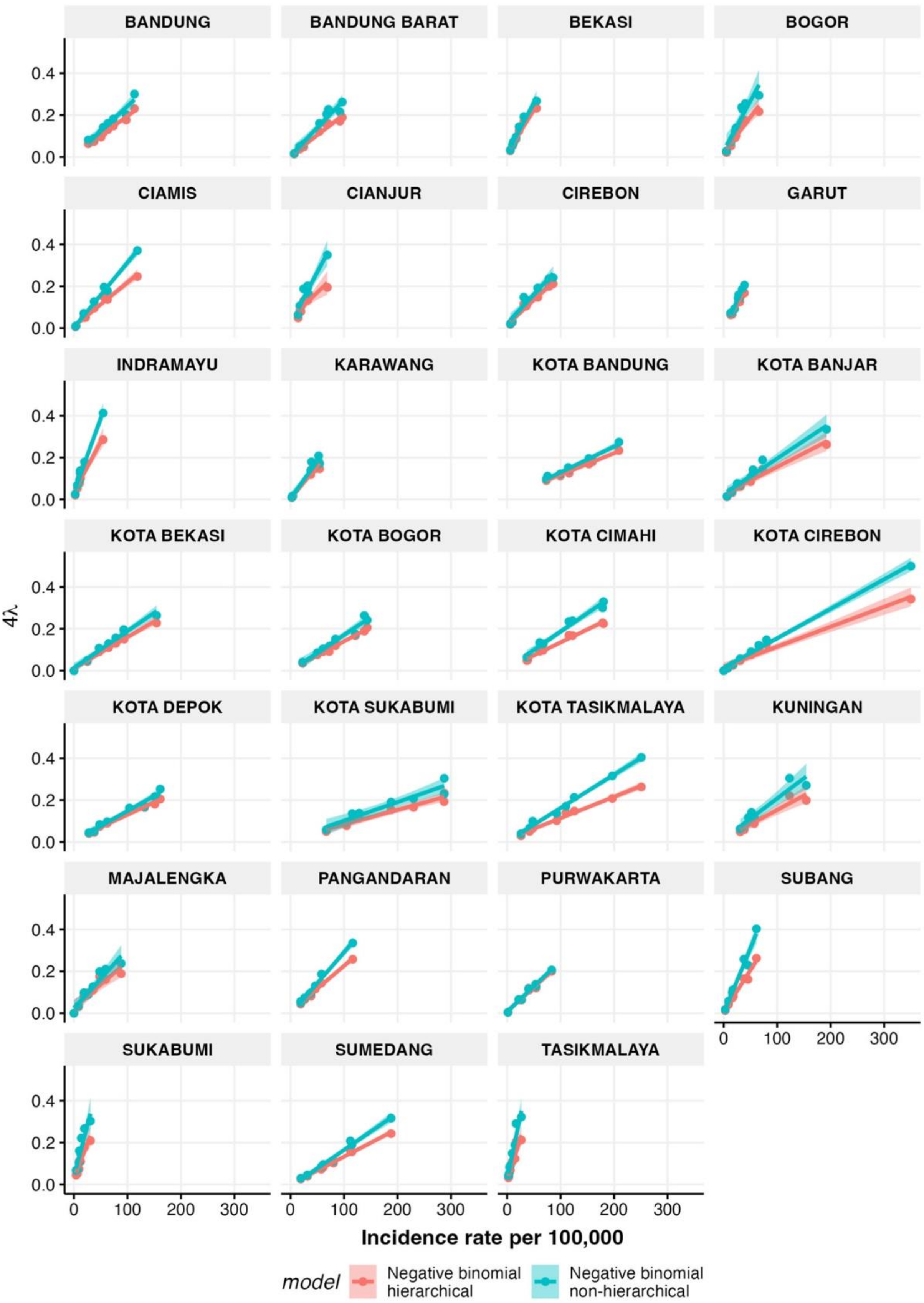


**Figure S3.** Scatter plots of district-level relationship between reported incidence rate and FOI estimates for both hierarchical and non-hierarchical models in West Java province.

Coloured lines denote the estimated linear regression lines between the two variables from each model estimates.

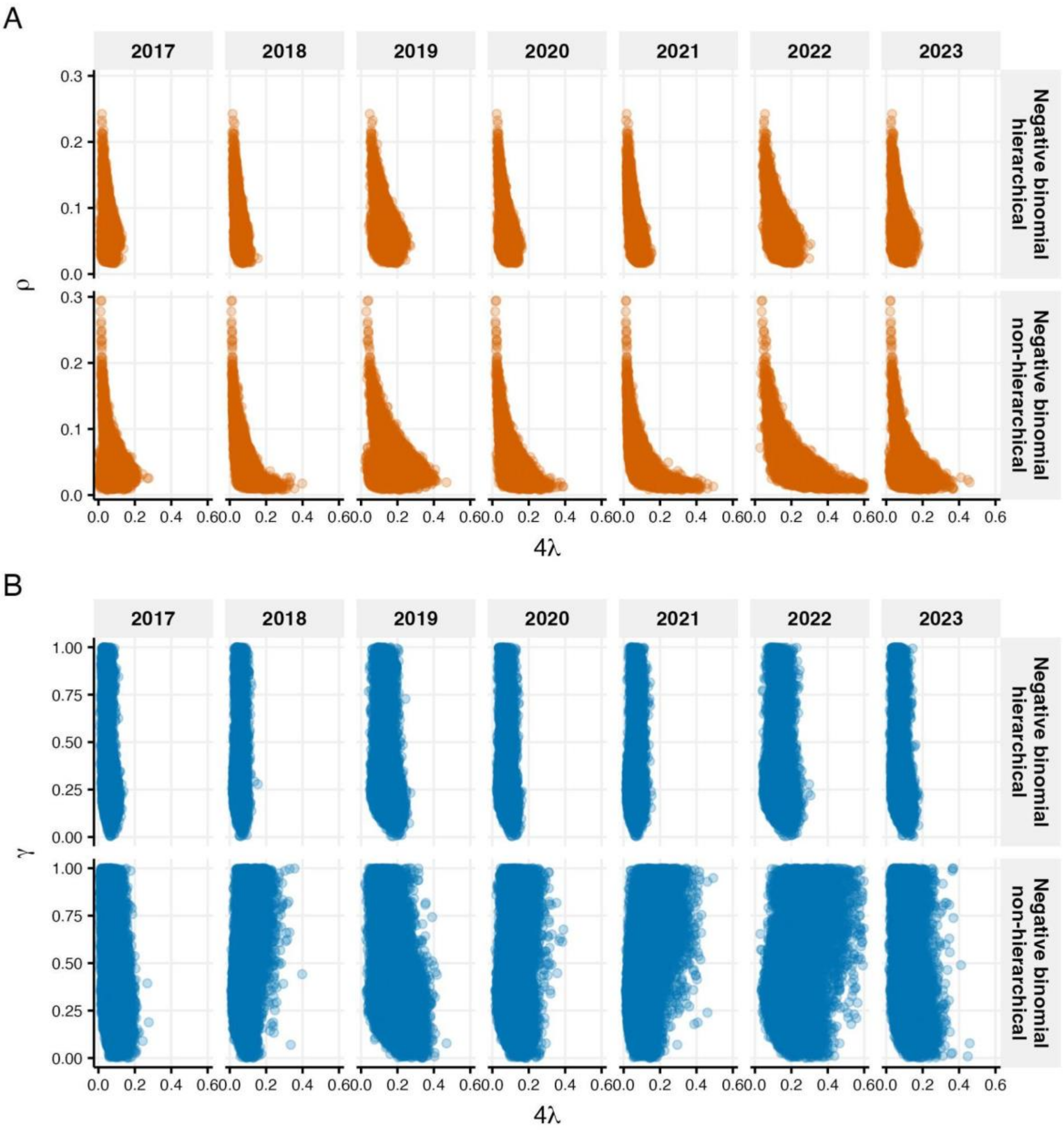


**Figure S4.** Correlations between FOI and $\rho$ (A) and $\gamma$ (B) for Jakarta using posterior samples

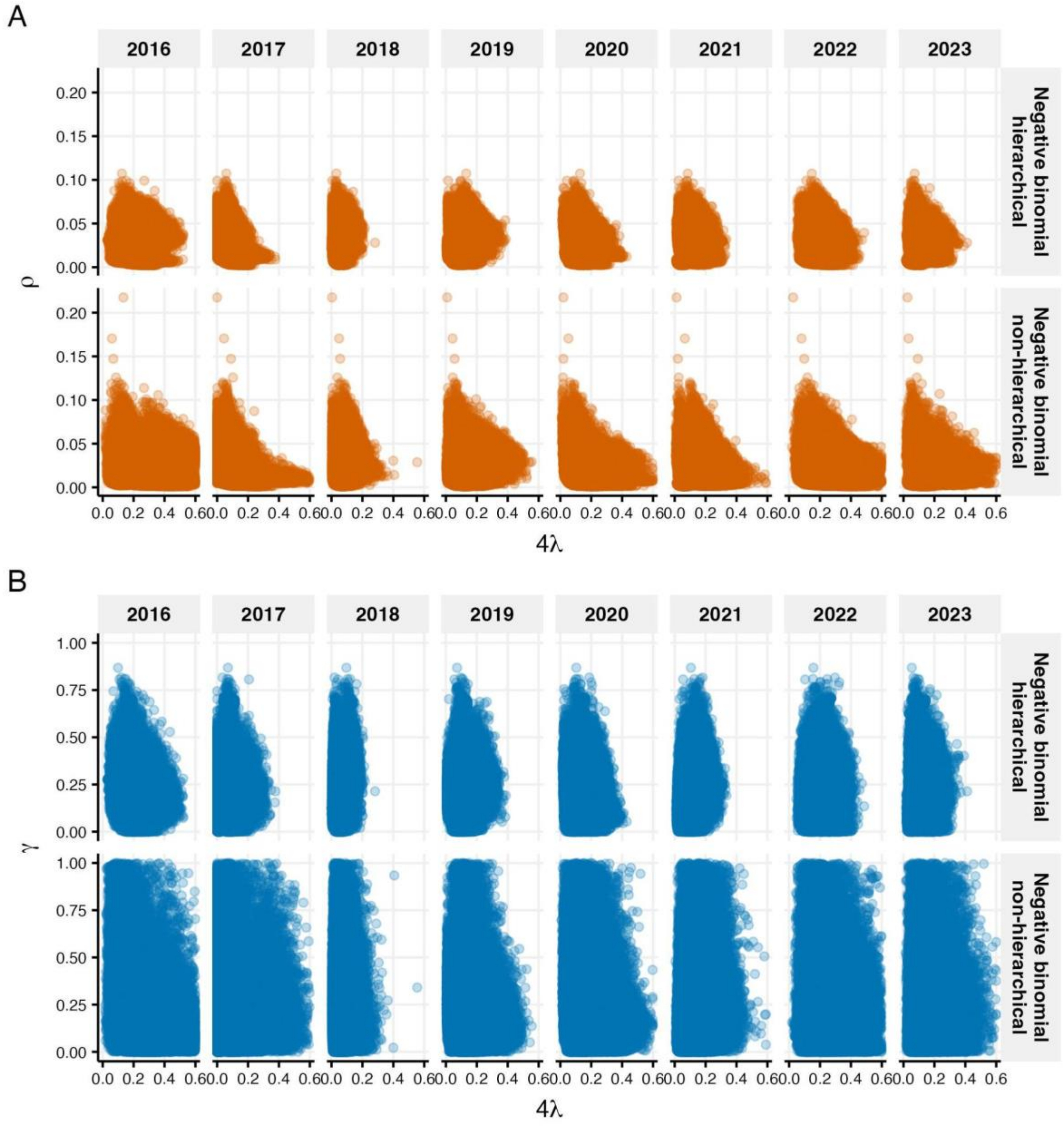


**Figure S5.** Correlations between FOI and $\rho$ (A) and $\gamma$ (B) for West Java using posterior samples

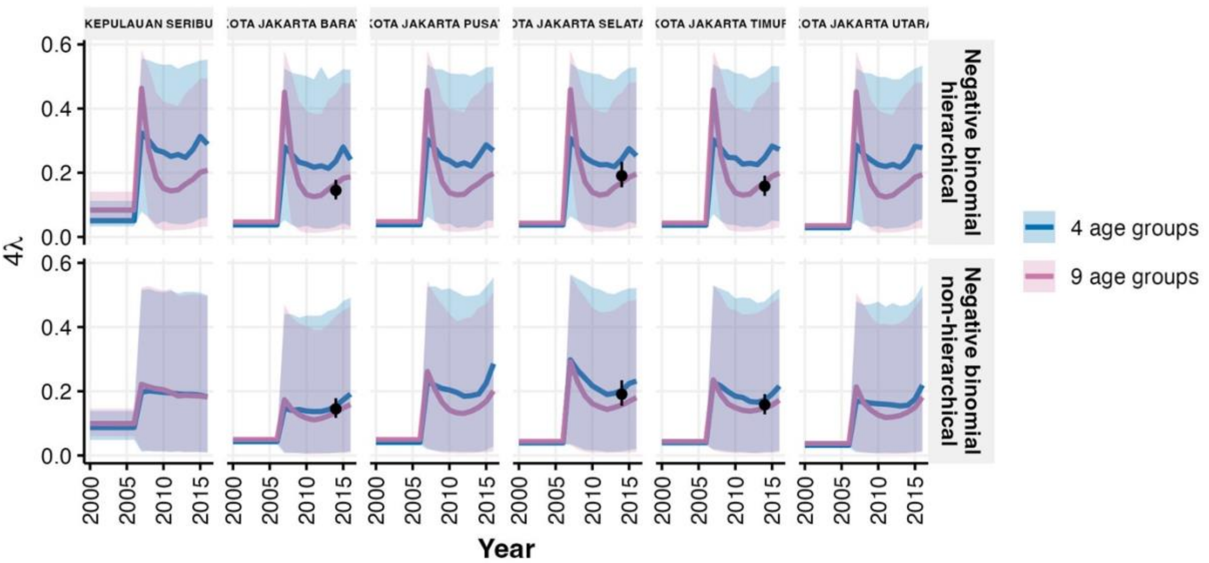


**Figure S6.** Sensitivity analysis comparing model estimates using four versus nine age groups for Jakarta districts. (A) Historical FOI estimates from hierarchical (top) and non-hierarchical (bottom) models, with black points showing 2014 seroprevalence-derived estimates.

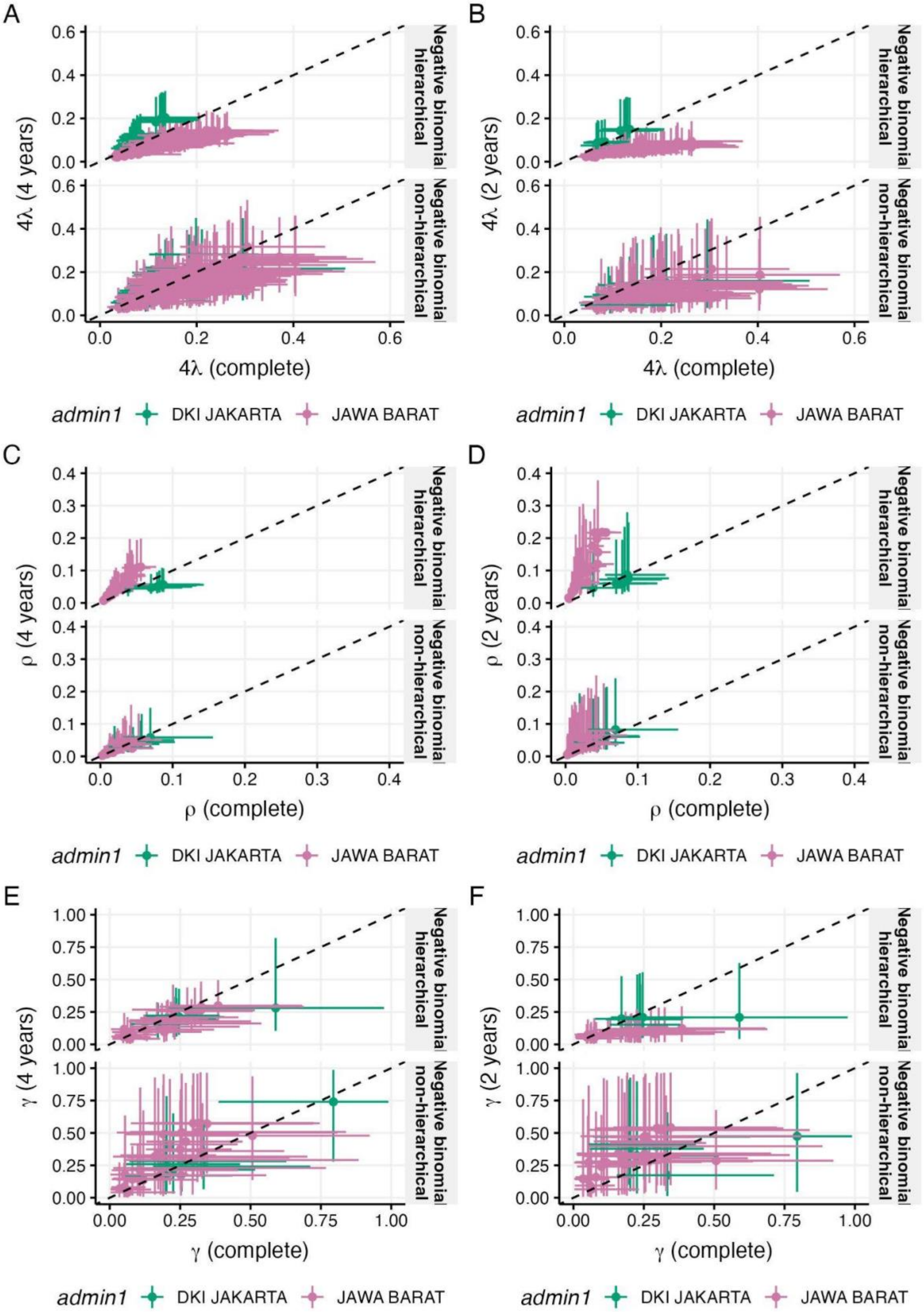

A
B
C
D
E
F
4λ (4 years)
4λ (2 years)
4λ (complete)
ρ (4 years)
ρ (2 years)
ρ (complete)
γ (4 years)
γ (2 years)
γ (complete)
Negative binomial hierarchical
Negative binomial non-hierarchical
admin1
DKI JAKARTA
JAWA BARAT

**Figure S7.** Sensitivity of model estimates with temporal length of data left 4 years and right 2 years

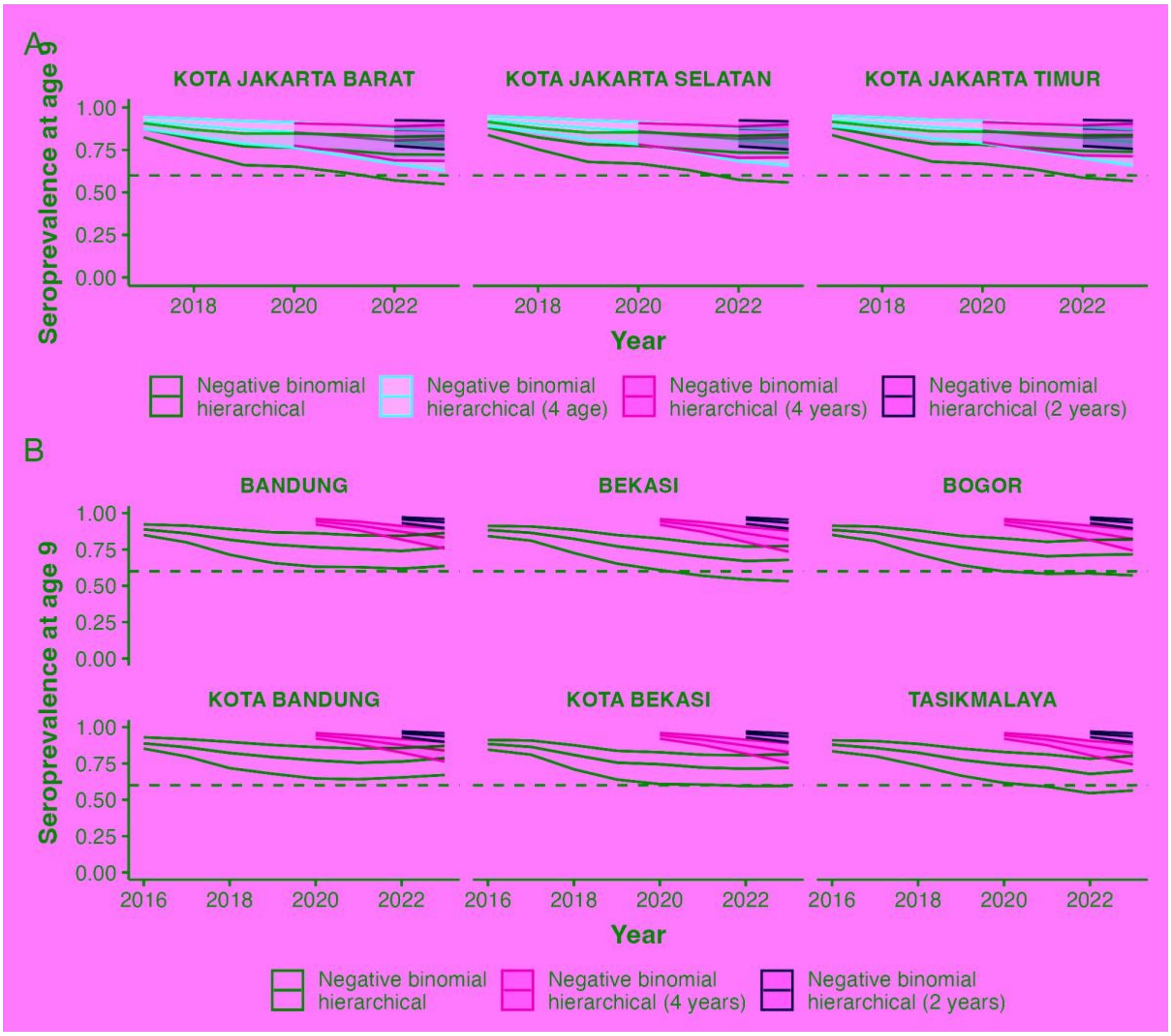


**Figure S8.** Estimates of the seroprevalence at age 9 years old hierarchical (A) selected Jakarta districts, (B) selected West Java districts

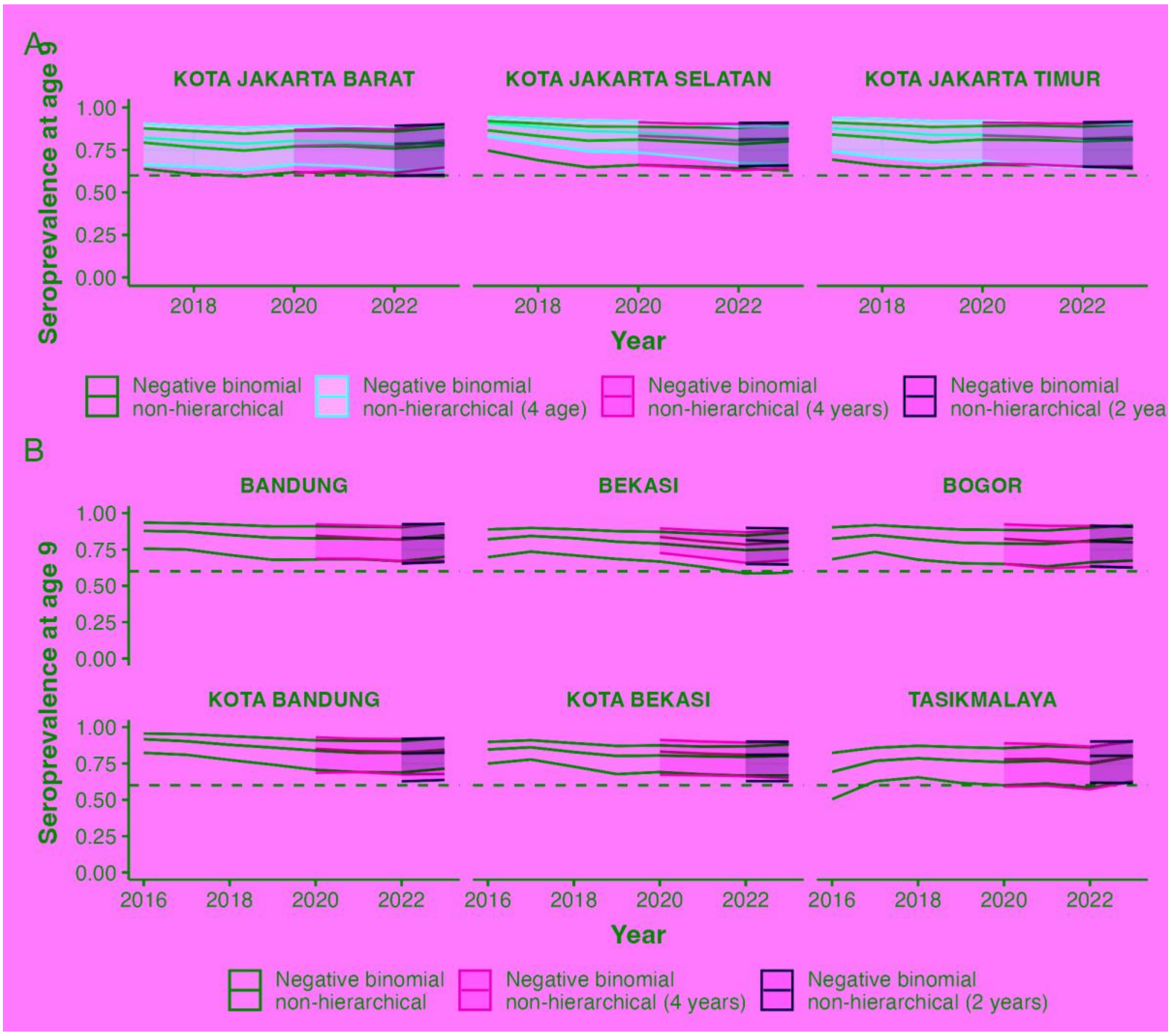


**Figure S9.** Estimates of the seroprevalence at age 9 years old non-hierarchical models (A) selected Jakarta districts, (B) selected West Java districts
